# Cervicovaginal inflammation and HIV target cell activation in adolescent girls and young women with *Chlamydia trachomatis* infections

**DOI:** 10.1101/2025.02.04.25321658

**Authors:** Smritee Dabee, Shaun Barnabas, Brian Kullin, Bart Versteeg, Nonhlanhla Mkhize, Etienne Muller, Venessa Maseko, David A. Lewis, Janan Dietrich, Glenda Gray, Katherine Gill, Linda-Gail Bekker, Musalula Sinkala, Darren P. Martin, Sylvia M. Bruisten, Heather B. Jaspan, Jo-Ann S. Passmore

## Abstract

**Background:** Adolescent girls and young women (AGYW) in sub-Saharan Africa are disproportionately affected by *Chlamydia trachomatis* (CT) infections, a leading cause of pelvic inflammatory disease (PID), infertility, and increased HIV susceptibility. CT-induced genital inflammation disrupts mucosal barriers and activates HIV target cells, compounding reproductive and immunological risks. We investigated the impact of CT infection in AGYW from two South African regions, focusing on genital inflammation and immune activation.

**Methods:** An observational cohort of 298 sexually active AGYW (16–22 years) from Cape Town and Johannesburg was enrolled. Participants in Johannesburg attended a single study visit, while those in Cape Town were followed longitudinally for up to three visits over 6–8 months. Cytokine profiling of cervicovaginal samples assessed inflammatory responses, while cervical cytobrush-derived CD4+ T cells were analyzed for CD38, Ki67 and CCR5 expression. CT *ompA* genotyping examined strain diversity.

**Results:** CT prevalence was 2.4 times higher in Cape Town (41.6%) than in Johannesburg (17.4%). Infection with CT led to an upregulation of inflammatory cytokines, including IL-1β, IL-6, and TNF-α, with a stronger inflammatory response observed in Johannesburg. Genotypic analysis revealed regional variation: genovar D, predominant in Cape Town, was associated with lower IFN-γ concentrations, whereas genovar E, more common in Johannesburg, was associated with higher IFN-γ levels. AGYW who experienced CT infection across multiple study visits did not exhibit elevated genital tract cytokine levels compared to those with a single infection. However, they did show an increase in activated and proliferating cervical CD4+ T cells, which may contribute to heightened HIV susceptibility.

**Conclusions:** CT genotypic diversity and regional variation are associated with differences in immune responses in AGYW, alongside important contributions from host immune history.

These findings support the need for integrated sexual health services and inform future regionally relevant vaccine strategies.

## Introduction

Young women in sub-Saharan Africa face disproportionately high risks of HIV infection, driven by both biological and socio-behavioural vulnerabilities (1). Genital inflammation is a critical factor that increases the risk of HIV acquisition (2). Sexually transmitted infections (STIs), including *Chlamydia trachomatis* (CT) are major drivers of genital inflammation. CT infections are frequently asymptomatic in women, leading to under-treatment despite associations with pelvic inflammatory disease (PID), infertility, ectopic pregnancy, and increased HIV risk due to subclinical inflammation (3).

CT infection induces genital cytokine responses that play a key role in pathogenesis (4). Pro-inflammatory mediators such as IL-1β, IL-6, and IP-10 can also exacerbate tissue damage and increase HIV susceptibility through mucosal barrier disruption and CD4+ T cell activation (2,5,6). Persistent CT infection can further amplify these inflammatory responses, contributing to immune dysregulation and impaired clearance (6).

CT exhibits substantial genetic diversity that influences transmission dynamics, tissue tropism, and host immune responses. Urogenital CT infections in women, including AGYW in sub-Saharan Africa, are dominated by a limited number of *ompA*-defined genotypes, particularly E, D, and F, with genotypes J, G, and K contributing smaller but consistent proportions across regions (7–8). Variation within and between *ompA* genotypes has been associated with differences in immune activation, persistence, and clinical outcomes, underscoring the relevance of characterizing circulating CT genotypes in high-burden African AGYW populations (9).

AGYW are particularly vulnerable to CT infection due to socio-behavioral, anatomical, and hormonal factors. The adolescent cervix has a larger area of ectropion (10), exposing columnar epithelium that is more permissive to bacterial adhesion and invasion than the stratified squamous epithelium in mature women. Together with delayed diagnosis and reinfection, these factors increase the risk of prolonged inflammation and reproductive complications in AGYW (11).

Given these challenges, it is critical to investigate the complex interplay between CT genotypes, host inflammatory responses, and mucosal immune activation in AGYW. We examined how CT genotypes influence genital inflammation and immune activation in AGYW from two South African regions.

## Materials and Methods

### Cohort description

Between November 2013 and February 2015, AGYW (16–22 years) were recruited from Cape Town (n=149) and Johannesburg (n=149) through the Desmond Tutu HIV Centre (DTHC) and the Perinatal HIV Research Unit (PHRU), respectively (12). The study was cross-sectional in Johannesburg while the cohort from Cape Town arm were followed longitudinally for up to three visits, using a targeted enrolment strategy with predefined sample sizes (Barnabas et al., 2018). For the longitudinal cohort, study visits were 3 monthly unless participants were using Net-EN, in which case it was 2 monthly. AGYW using the injectable contraceptives (DMPA or Net-EN) had visits scheduled two weeks post-injection, while non-users were scheduled during the luteal phase (days 14–28) of their menstrual cycle. Participants were excluded if they were HIV-positive, pregnant, sexually inactive, douched or used spermicides in the past two days, or took antibiotics in the past two weeks. Research Ethics Committees of the Universities of Cape Town and Witwatersrand approved the study (#267/2013 and #M130745, respectively). Written informed consent was obtained from participants 18+ years and assent was provided by those <18 years, with written consent from a parent/legal guardian.

### Sample collection

At enrolment, participants completed a socio-demographic questionnaire (12). At each visit, cervicovaginal fluid was collected using a Softcup® menstrual cup, inserted for one hour. During the clinical speculum exam, a Dacron vulvovaginal swab was taken for STI testing, a FLOQSwab™ collected lateral vaginal wall samples for BV screening (Nugent scoring; 13), and cervical mononuclear cells were obtained using a Digene® endocervical cytobrush.

### Testing for STIs and BV

Vulvovaginal samples were tested for discharge-causing STIs (CT*, Neisseria gonorrhoeae* [NG]*, Trichomonas vaginalis* [TV]*, Mycoplasma genitalium* [MG]), as well as ulcer-causing STIs (HSV-1, HSV-2, *Haemophilus ducreyi* [HD]*, Treponema pallidum)*, using real-time multiplex PCRs (14). BV was diagnosed by Nugent score (13). AGYW positive for CT were treated with Azithromycin (1g orally; single dose), those with NG given Ceftriaxone (250mg intramuscular injection; single dose), and those positive for BV (Nugent 7-10) or TV were given Metronidazole (400mg orally, twice daily; 7 days), according to South African national guidelines at the time. AGYW with CT or NG were given partner notification letters and referred to the local Department of Health clinic.

### Chlamydia *ompA* genotyping

*OmpA* genovar diversity of CT was analyzed by MLST genotyping (8) (15). Sequence data were analysed using the CT database hosted on PubMLST (https://pubmlst.org/bigsdb?db=pubmlst_chlamydiales_seqdef). The *ompA* genovars were assigned by comparison to an offline reference database described previously (8) (15).

### Measurement of cytokine concentrations

Softcups were collected (in 50ml Falcon tubes), maintained at 4°C after collection (as previously described; 16, 17), and processed at a site-adjacent laboratory within 4 hours for storage at-80°C. Processing and elution of cervicovaginal fluid included inverting the softcup membrane using a sterile wooden disposable spatula in a BSL2 hood and centrifuging (1500 rpm, 10 minutes). The volume of secreted fluid was calculated, as previously described (17). The softcup was discarded. Assuming 1g=1mL, secretions were diluted 10 times with PBS and stored immediately at-80oC in aliquots. All softcup samples were transported to the UCT laboratory on dry ice, aliquots thawed in batches, and cytokines measured by Luminex using the same lot number of kits (16). Prior to Luminex, all samples were filtered by centrifugation (1950g, 10 min, 4°C) using SPIN-X® 0.2μm filters. Two different kits were used: the Human Cytokine Group I 27-plex (FGF-basic, Eotaxin, G-CSF, GM-CSF, IFN-γ, IL-1β, IL-1RA, IL-2, IL-4, IL-5, IL-6, IL-7, IL-8, IL-9, IL-10, IL-12(p70), IL-13, IL-15, IL-17, IP-10, MCP-1, MIP-1α, MIP-1β, PDGF-BB, RANTES, TNF-α, and VEGF) and the Human Cytokine 21-plex kit (IL-1α, IL-2RA, IL-3, IL-12(p40), IL-16, IL-18, CTACK, GRO-α, HGF, IFN-α2, LIF, MCP-3, M-CSF, MIF, MIG, NGF, SCF, SCGF-β, SDF-1α, TNF-β, and TRAIL) (Bio-Rad). Experiments followed manufacturer instructions. Samples from both sites were randomly allocated on the different assay plates, with the same ten samples included in all plates: Inter-and intra-plate correlations (Spearman correlations) between these ten samples were carried out for each cytokine amongst all the plates. IL-15 was excluded as the values did not pass the QC. The cytokines IL-2, IL-5 and RANTES were not included in the analysis as 76.0%, 64.2% and 52.6% of the values respectively were below detectable range. For the rest, detection limits ranged from 1-307 pg/ml, and concentrations were determined using 5PL regression. Values below detection limits were recorded as half the lowest measured concentration.

### Flow cytometry

CMC samples from Cape Town participants were collected and analyzed via flow cytometry at each visit. Cervical cytobrush samples were processed as previously described (16), with the cell pellet resuspended in PBS, transferred to a 96-well plate, centrifuged at 300g for 3 minutes at 4°C, and washed before staining. An eight-color flow cytometry panel was used to assess CCR5 (APC; BD), activation markers CD38 (PE-Cy7; eBioscience) and HLA-DR (PE; BD), and proliferation marker Ki67 (FITC; BD) in CD4+CD3+ (APC-H7; BD) and CD8+CD3+ (QDot605, Invitrogen) T cells. CD8+ T cells is not shown. A dump channel (LIVE/DEAD® Fixable Violet Dead Cell Stain [Invitrogen], CD14 [PacBlue, BD], CD19 [Pac Blue, Invitrogen]) excluded dead cells, B-cells, and monocytes, respectively. Cells were first incubated with the live/dead marker for 20 minutes at room temperature, washed twice, and stained with extracellular markers. After 30-minute ice incubation, cells were washed, fixed and permeabilized using Cytofix/Cytoperm (BD). Intracellular markers CD3 and Ki67 were added, and cells incubated for 30 minutes on ice. The pellet was resuspended in BD CellFIX^TM^, transferred to a FACS tube, wrapped in foil, and stored at 4°C until acquisition on the BD LSRFortessa^TM^. Data were acquired within 48 hours, with compensation tubes prepared for each run. FlowJo (v10.0.8; Treestar) was used for analysis, with gating based on FMO controls.

## Statistical analysis

Statistical analyses and linear regressions were carried out using STATA 12.0 and R v3.6.0. Graphs and figures were generated using Prism 6 and R. Unpaired and paired continuous variables were compared using the Mann–Whitney U and Wilcoxon signed-rank tests, respectively, and categorical variables using two-sided chi-squared_tests. Analyses were adjusted for multiple comparisons using false discovery rate step down procedures.

## Results

A total of 298 sexually active AGYW (median age: 18 years; IQR: 17–20) from Cape Town and Johannesburg were included (12) (16). Johannesburg participants had a single-visit study, while Cape Town participants were followed over three visits spanning 6–8 months. Contraceptive use differed by site: in Johannesburg, 63.8% (77/121) of AGYW reported condom use alone, whereas in Cape Town 70.5% (105/149) used injectable hormonal contraception (DMPA or Net-EN). CT was the most prevalent STI (29.5%, 88/298), with CT prevalence being significantly higher in Cape Town (41.6%, 62/149; 95% CI: 33.6–49.9%) than in Johannesburg (17.4%, 26/149; 95% CI: 11.8–24.3%; p<0.0001). Among those with CT, 71.6% (63/88) had another STI or BV.

### Impact of CT infection on genital inflammation in AGYW

The genital cytokine responses to CT infection in AGYW at baseline, were assessed by multivariate linear regressions, adjusted for contraceptive use, co-infections with STIs and BV (Figure 1A). AGYW infected with CT exhibited significant upregulation of cytokines across all functional classes compared to uninfected individuals. These included IL-1β (β-coefficient=0.36; p=0.022), IL-6 (β=0.21; p=0.038), TNF-α (β=0.23; p=0.015), IP-10 (β=0.29; p=0.035), MIG (β=0.27; p<0.0001), G-CSF (β=0.28; p=0.018), HGF (β=0.27; p=0.011), SCF (β=0.13; p=0.034), IFN-γ (β=0.17; p=0.025), and IL-4 (β=0.17; p=0.015), after adjusting for multiple comparisons. A focused analysis on cytokines implicated in CT pathogenesis (IL-1α, IL-1β, IL-6, IL-8, TNF-α, TNF-β) and protection (IFN-γ), including AGYW with CT infection alone (n=26; no co-infections or BV) compared to uninfected controls (n=62; no STIs or BV) (Figure 1B), further confirmed that CT infection significantly elevated concentrations of inflammatory cytokines, including IL-1α (p=0.03), IL-6 (p=0.01), IL-8 (p=0.05), IP-10 (p=0.002), and TNF-α (p=0.01). Although protective IFN-γ levels were higher in CT cases compared to controls, this was not significant.

**Figure 1.**
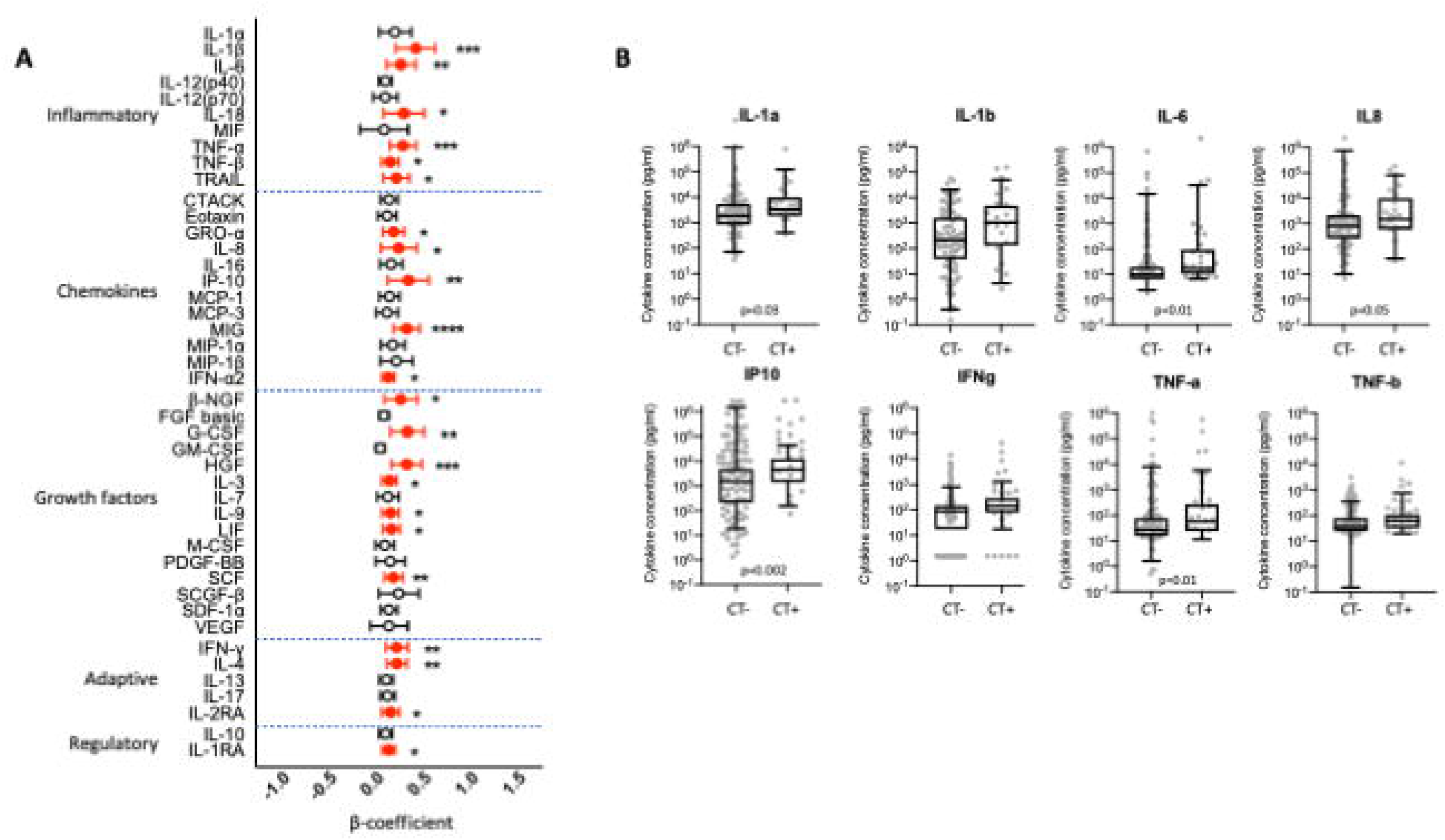
Impact of CT infection on cervicovaginal cytokine profiles in South African AGYW at baseline (only visit 1 for Cape Town AGYW). (A) Multivariate linear regression analysis illustrating associations between cervicovaginal cytokine levels and CT infection status, adjusted for participant age, hormonal contraceptive use, other co-infections, BV, and semen exposure. Each association is represented by a β-coefficient, with error bars indicating the 95% confidence intervals. Statistically significant associations (p ≤ 0.05 after correction for multiple comparisons) are denoted with filled symbols. (B) Concentrations of cervicovaginal cytokines associated with inflammation (IL-1α, IL-1β, IL-6, IL-8, IP-10, TNF-α, TNF-β) and protection (IFN-γ) are compared between AGYW with CT infections (n = 26; with no other STIs or BV) and those without CT (n = 62; no STIs or BV). Group comparisons were performed using Mann-Whitney U-tests with statistical significance set at p ≤ 0.05 after adjustment for multiple comparisons. For women in Cape Town for which multiple visits were available, only the baseline visit was included (no repeated measures).

### Chlamydia diversity and associated inflammation by study site

CT infection was more inflammatory in Johannesburg than in Cape Town at baseline (Figure 2A). In Cape Town, CT was linked to MIG upregulation (not significant after multiple comparisons), while in Johannesburg, it was associated with increased concentrations of 34/44 cytokines. Among AGYW with CT alone (no other STIs or BV), inflammatory cytokines tended to be higher in Johannesburg (n=4 CT+; no other STIs/BV) than Cape Town (n=22 CT+; no other STIs/BV), although numbers were small (Figure 2B).

**Fig 2.**
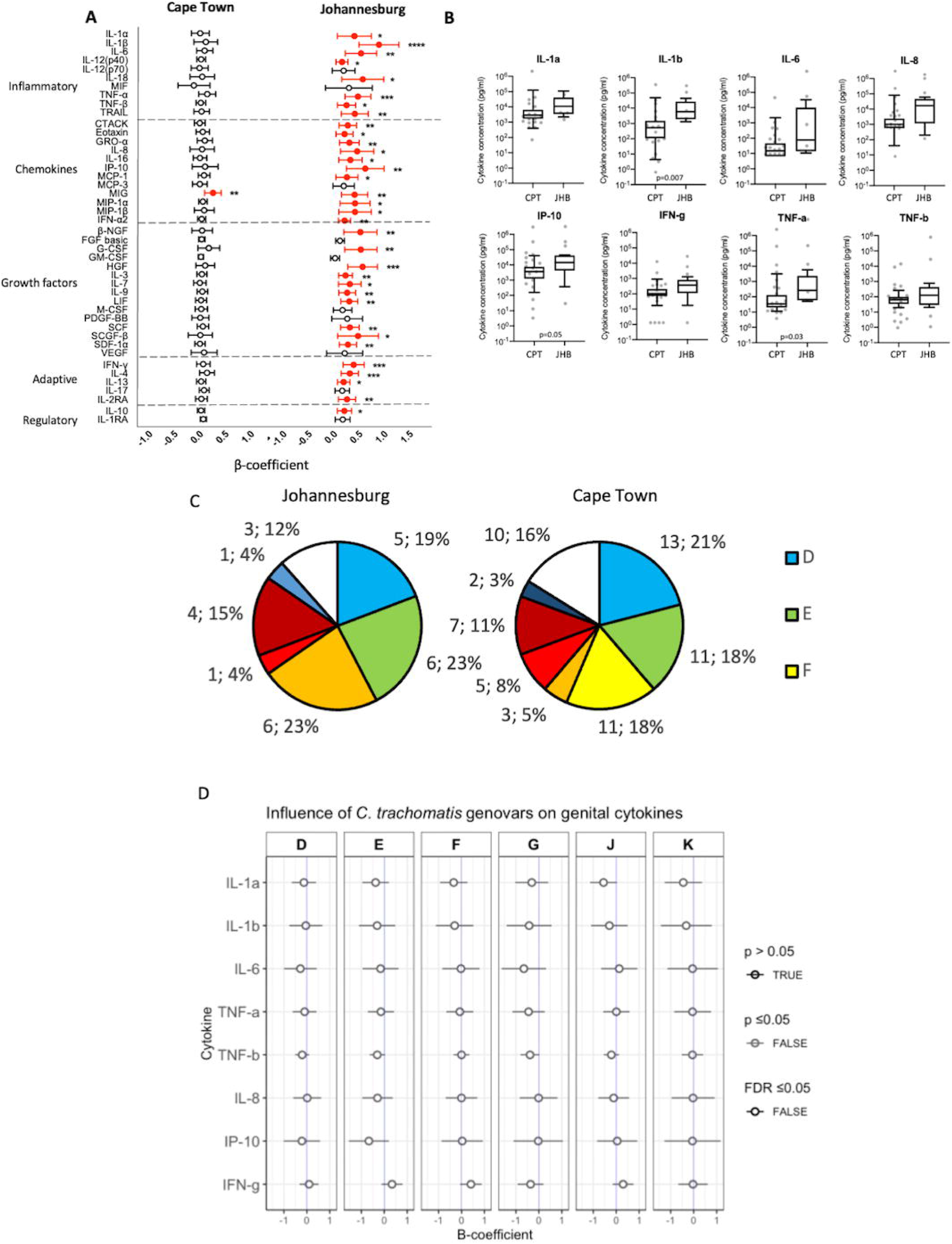
Investigating the impact of CT genovar on inflammatory responses in AGYW from Cape Town and Johannesburg at baseline. (A) Multivariate linear regression analysis comparing cervicovaginal cytokine levels associated with CT infection in AGYW from Cape Town (left panel) and Johannesburg (right panel), adjusted for participant age, hormonal contraceptive use, other co-infections, BV, and semen exposure. Associations are represented by β-coefficients, with error bars indicating 95% confidence intervals. Statistically significant associations (p ≤ 0.05 after correction for multiple comparisons) are marked with filled symbols. (B) Cervicovaginal cytokine concentrations associated with inflammation (IL-1α, IL-1β, IL-6, IL-8, IP-10, TNF-α, TNF-β) and protection (IFN-γ) are compared between AGYW with CT infections in Cape Town (CPT) and Johannesburg (JHB) (n = 26; with no other STIs or BV). Group comparisons were conducted using Mann-Whitney U-tests, with significance defined as p ≤ 0.05 after adjustment for multiple comparisons. For women in Cape Town for which multiple visits were available, only the baseline visit was included (no repeated measures). (C) Distribution (n, percentage) of CT ompA genotypes in AGYW from Johannesburg (left pie chart) and Cape Town (right pie chart). Genovar D is represented in blue, E in green, F in yellow, G in orange, I in bright red, J in dark red, K in purple, and unknown genotypes in white. (D) Multivariate linear regression analysis of associations between cervicovaginal cytokine levels and CT ompA genovars (D to K), adjusted for participant age, hormonal contraceptive use, other co-infections, BV, and semen exposure. Associations are represented by β-coefficients with 95% confidence intervals. None of the associations were Statistically significant (p ≤ 0.05) after correction for multiple comparisons.

To assess whether CT strain diversity contributed to differences in inflammatory responses, *ompA* genotyping was performed on 128 samples with sufficient DNA (Figure 2C), including 20/26 cases from Johannesburg and 108/121 cases from Cape Town (spanning all three study visits). Eighteen samples were excluded due to low DNA quantity. The distribution of *ompA* genovars did not differ significantly between sites (p = 0.1; χ² test), and globally predominant urogenital genotypes were represented at both sites. In Cape Town, genovar D was the most prevalent (21%, 13/62), followed by genovars E (18%, 11/62) and F (18%, 11/62), while genovar G was detected in 5% of cases (3/62). In contrast, in Johannesburg, genovars E and G were most common (23% each, 6/26), while genovar D was detected in 19% (5/26) of cases and genovar F was not detected. Less common genovars (I, J, K) were observed at low frequencies at both sites. No significant associations were observed between individual CT genovars and inflammatory cytokines, after adjustment for multiple comparisons (Figure 2D). However, the limited number of CT-positive samples, particularly within specific genovars and at the Johannesburg site, constrained statistical power and precluded site-specific comparisons of genovar-associated inflammatory profiles.

### Inflammation associated with persistent CT infections

In Cape Town, where AGYW were followed longitudinally, 33.9% (43/127) were infected with CT at visit 2, and 18.2% (16/88) were infected at visit 3 (Table 1). Among participants with repeat CT detection, genotyping indicated that the infecting genovar was most often the same across visits: 10 women were CT-positive at all three study visits, of whom 7/10 harbored the same genovar at each visit; 10 were CT-positive at visits 1 and 2 only, all of whom (10/10) had the same genovar at both visits; and 5 were CT-positive at visits 1 and 3 but not visit 2, of whom 4/5 had the same genovar at both positive visits. These findings suggest that repeated detection of CT in this cohort was more frequently associated with persistence or re-exposure to the same strain, which may reflect ongoing exposure from the same sexual partner or incomplete clearance, rather than acquisition of a new genotype. To investigate whether persistent or repeat CT infections were associated with increased inflammation, cervicovaginal cytokine concentrations were compared among women infected at one, two, or three visits (Figure 3). The analysis revealed no significant differences in the concentrations of any measured cytokines across these groups, regardless of the duration of infection.

**Figure 3.**
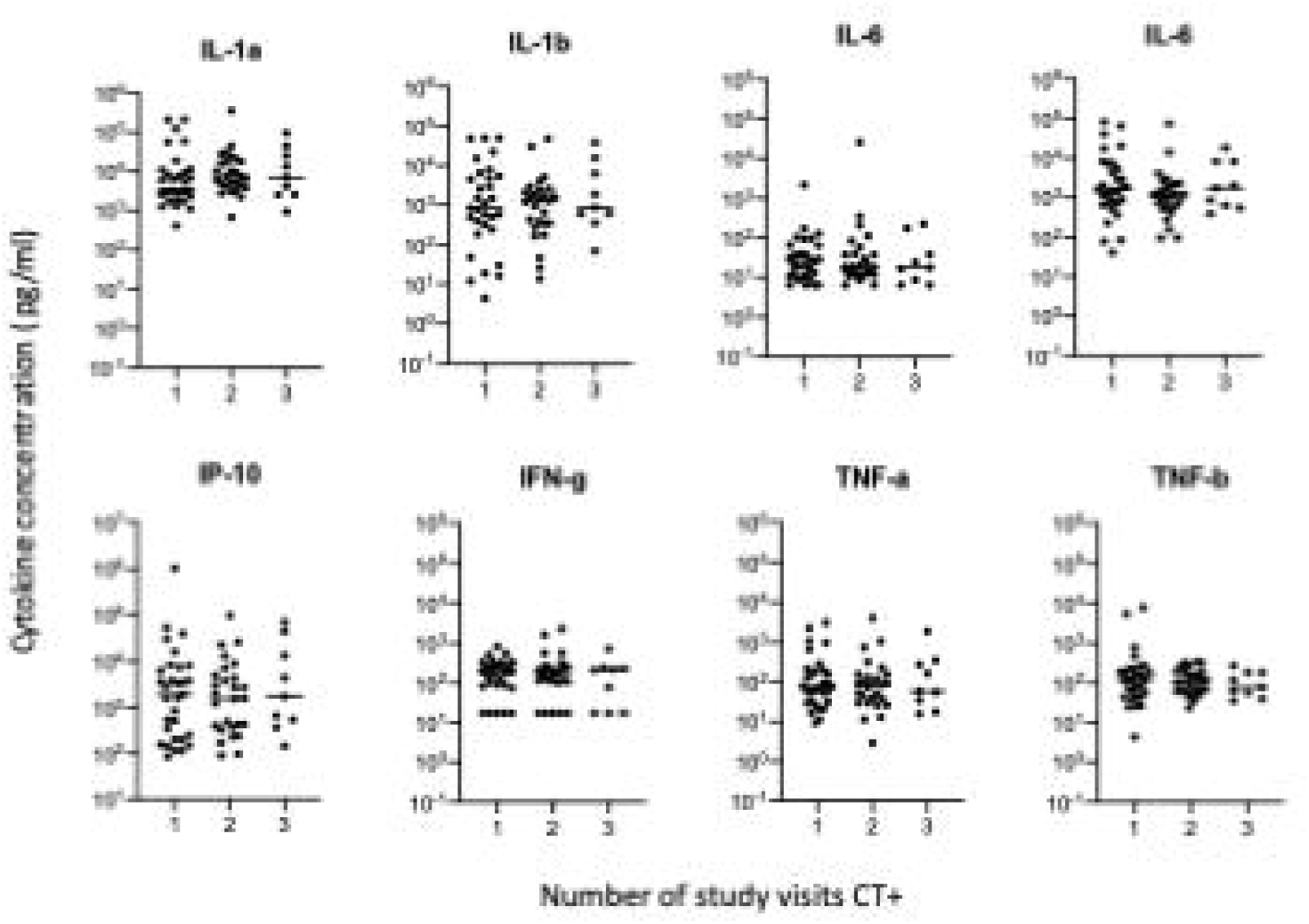
Cervicovaginal cytokine concentrations in AGYW infected with CT over one, two, or three study visits. Each dot represents the cytokine response of an individual participant, with black lines indicating the median values for each group. Group comparisons were conducted using Mann-Whitney U-tests, with statistical significance defined as p ≤ 0.05.

**Table 1.**
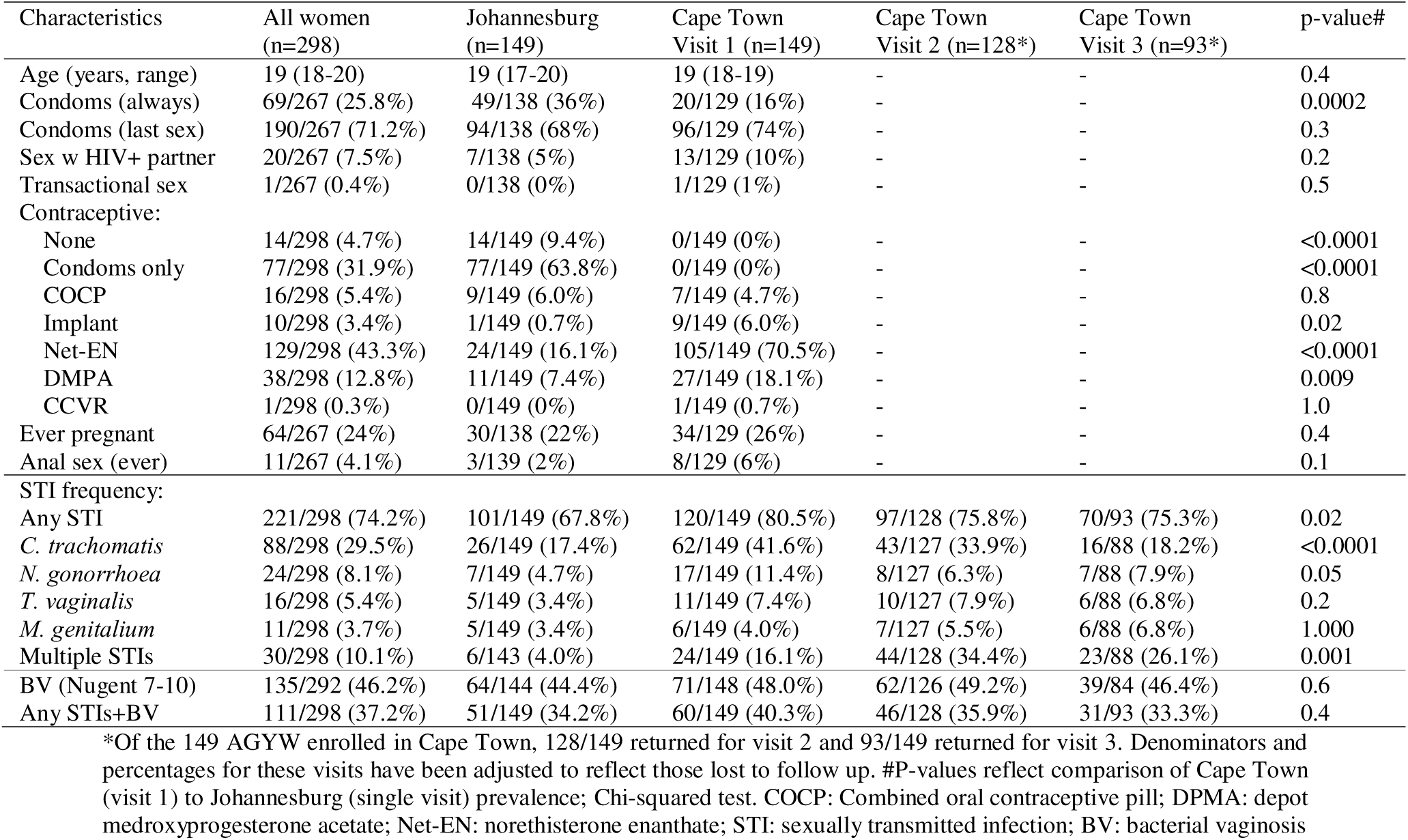
Characteristics for study participants.

### Genital tract HIV target-cell activation associated with CT infections

Cervical cytobrush-derived CD3+CD4+ T cells were used to assess the expression of CCR5 (HIV entry receptor on CD4+ cells), CD38 (a marker of cellular activation), and Ki67 (a marker of T-cell proliferation) (Cape Town only). Figure 4A shows a representative figure of the gating strategy. The frequency of activated cervical CD38^+^ cells among CD3+CD4^+^ T cells was significantly higher in CT cases compared to controls (p=0.05; Figure 4B, top panel). However, there were no significant differences in the frequencies of CD3+CD4^+^ T cells or those expressing CCR5 or Ki67. As a technical observation, CT+ cytobrush specimens yielded higher total acquired live events under standardized acquisition settings (Figure 4B, bottom panel).

**Figure 4.**
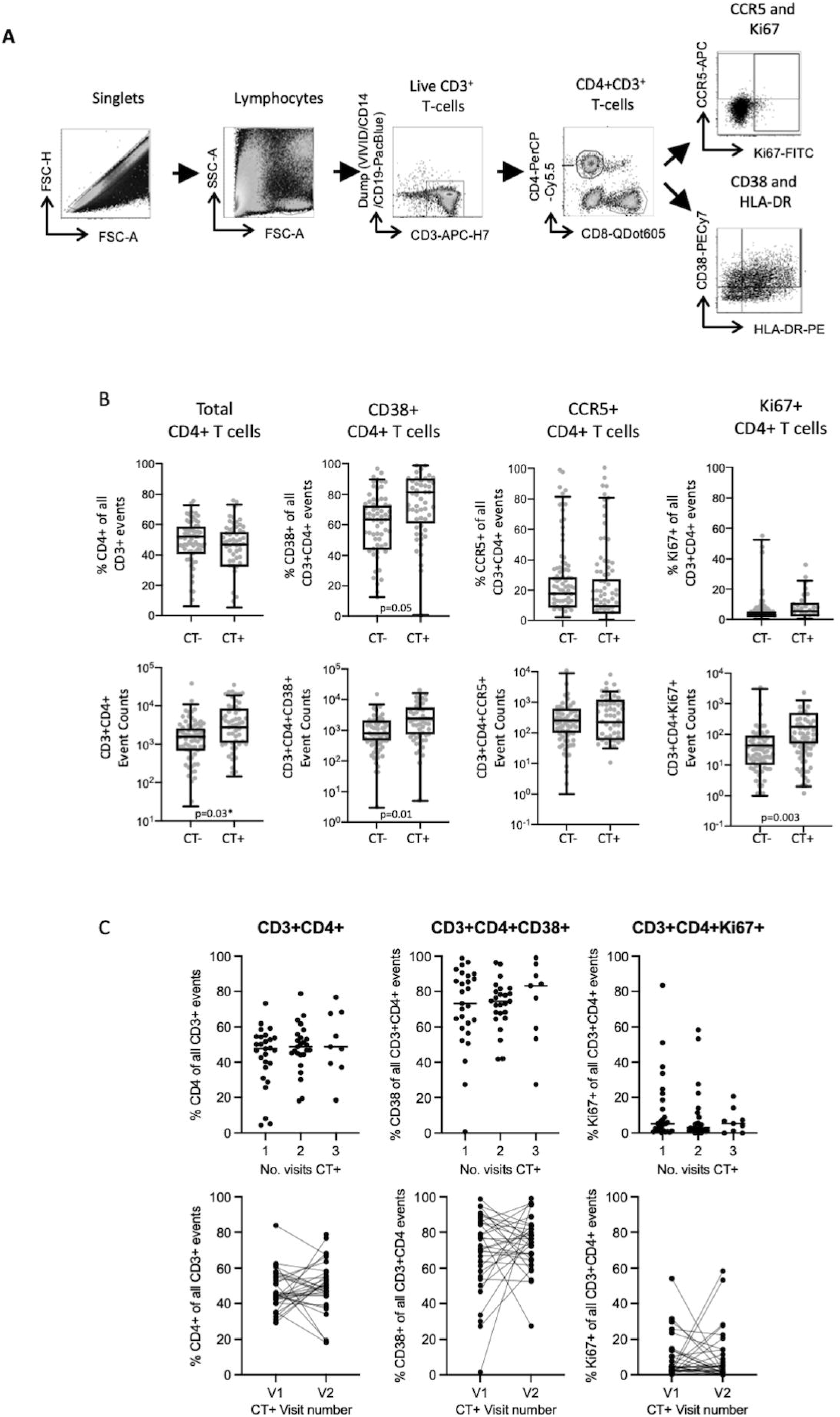
Impact of prevalent or persistent CT Infections on cervical cytobrush-derived CD4+ T cell activation. (A) Representative gating strategy for flow cytometry analysis of cervical cytobrush-derived T cells, focusing on live CD3+CD4+ cells expressing activation marker CD38+, proliferation marker Ki67+, or HIV co-receptor CCR5. (B) The frequency (top panel) and absolute number of events (bottom panel) are shown for total CD3+CD4+ T cells (far left), CD3+CD4+ T cells expressing CD38 (middle left), CCR5 (middle right), and Ki67 (far right). Data were collected at visit 1 (study entry) from N = 36 controls and N = 18 CT cases. (C) Longitudinal analysis of cervicovaginal CD3+CD4+ T cell activation marker expression is presented for AGYW with CT infections spanning one, two, or three study visits (top panel). Matched analyses for participants with repeat CT infections across multiple visits are shown in the lower panel. Statistical comparisons were performed using Mann-Whitney U-tests for unpaired analyses (B) and Wilcoxon Ranks test for paired data (C), with significance defined as p ≤ 0.05.

We further compared CD3+CD4^+^ T cell events in women infected with CT at one, two, or three study visits (Figure 4C). The frequency of activated or proliferating cervical CD3+CD4^+^ T cells did not differ significantly between women infected at multiple time points and those infected at a single visit. In a paired analysis of women infected at two study visits (n=32), there was no significant difference in the frequency of T cells measured at the second positive visit compared to the first positive visit, and frequencies did not correlate significantly (CD3+CD4+ R^2^=0.002 p=0.8; CD3+CD4+CD38+ R^2^=0.02 p=0.5; CD3+CD4+Ki67+ R^2^=0.02 p=0.5). This suggests that inflammation did not resolve or intensify with persistent or repeated infections (Figure 4C, lower panel).

## Discussion

Understanding how asymptomatic CT infection contributes to HIV susceptibility in AGYW remains an important challenge for prevention. Here, we examined the intersection between CT infection, genital inflammation, and HIV target cell availability across two South African settings, highlighting a complex interplay between pathogen diversity, host immune history, and local mucosal context. Rather than implicating a single explanatory factor, our findings indicate that inflammatory and cellular immune responses to CT reflect multiple determinants, including strain-level variation, prior exposure, and site-specific host or vaginal milieu factors (microbiome), helping to explain why inflammatory outcomes may differ across populations despite exposure to common CT lineages.

Genotyping revealed few differences in ompA genovar distribution between sites, with genovar D predominating in Cape Town and genovars E and G more frequent in Johannesburg. Despite this, AGYW with CT infection in Johannesburg exhibited a stronger overall inflammatory signature, even though genovar G—enriched at this site—was associated with lower cytokine concentrations. As this conclusion is based on the markers measured, site-level inflammatory differences are unlikely to be explained by genotype alone and may instead reflect additional unmeasured inflammatory or regulatory pathways and broader geographic, host, or vaginal milieu factors.

While variation in ompA genovars was explored, emerging evidence indicates that host immune history is likely a more dominant determinant of inflammation. In the same cohort, prior CT exposure and the quality of CT-specific CD4□ T cell responses strongly influenced genital immune profiles, with robust systemic Th1 responses inversely associated with mucosal inflammatory mediators (18). Women with untreated or recurrent CT infection showed increased genital T cell activation alongside impaired CT-specific T cell function, consistent with immune dysregulation following repeated exposure. Together, these findings suggest that adaptive immune history may outweigh strain-level variation, and that genovar associations should be regarded as hypothesis-generating rather than causal.

Strain-specific associations nonetheless highlight heterogeneity in CT immunogenicity. The association of genovar D with lower IFN-γ and genovar E with higher IFN-γ supports a role for pathogen genetic variation in shaping host responses. IFN-γ can drive CT into a persistent state via induction of indoleamine-2,3-dioxygenase (IDO), promoting immune evasion and chronic infection (19). Supporting this, Van Duynhoven et al. (20) reported greater lower abdominal pain among women infected with F or G group genotypes. Collectively, these findings may reflect variation in immunodominant T-cell epitopes within the major outer membrane protein (MOMP) (21), reinforcing the value of strain-level characterization for understanding CT pathogenesis and informing vaccine design.

This study has several limitations. Longitudinal data were only available from Cape Town, limiting analyses of repeat CT infections, and cellular activation measurements were similarly restricted. Partner treatment data were not systematically collected, and post-treatment cure testing was not performed. Longitudinal ompA genotyping in Cape Town showed that repeat CT detection usually involved the same genovar, suggesting persistence or re-exposure to the same strain, although reinfection from the same partner or incomplete clearance cannot be excluded. Genotyping was based on ompA sequencing rather than whole-genome sequencing, which may not capture within-genotype diversity (21) or recombination events (22). The small number of CT-positive participants in Johannesburg may also limit generalizability of site-specific genotype associations.

The high prevalence of CT-associated inflammation in AGYW highlights the need for improved STI screening and integrated sexual and reproductive health services in high HIV-burden settings (23). Regional differences in CT immunogenicity support tailored prevention approaches, including regionally informed vaccine strategies. In conclusion, our findings show that CT-associated genital immune responses in AGYW reflect the interplay of strain diversity, regional context, and host immune history.

## Data Availability

All data produced in the present study are available upon reasonable request to the authors

## Acknowledgements

We would like to thank the WISH study team, especially Ms Penelope Ngcobo (participant recruitment at DTHF), Sr Janine Nixon (sample collection), and all the young women enrolled in the study. We also thank Ms Michelle Himschoot from GGD Amsterdam, who assisted with the MLST lab work. This research was funded by the EDCTP Strategic Primer grant (SP.2011.41304.038; PI J. Passmore, UCT) and a self-initiated grant from the South African Medical Research Council (2017-2020; PI J. Passmore, UCT).

